# Effect of the conventional gait model 2 variants on lower-limb kinematics in individuals with cerebral palsy

**DOI:** 10.64898/2026.01.12.26343924

**Authors:** C Dussault-Picard, M Sangeux, S Armand, M Fonseca, F Leboeuf

**Affiliations:** Laboratory of Motion analysis, Physical Medicine and Rehabilitation, University Hospital of Nantes, France; Centre for clinical motion analysis, University Children’s Hospital Basel, Basel, Switzerland; Kinesiology Laboratory, Geneva University Hospitals and University of Geneva, Geneva, Switzerland; Centre of Research on skeletal Muscle and Movement, Geneva University Hospitals and University of Geneva, Geneva, Switzerland; Nantes Université, CHU de Nantes, Movement - Interactions - Performance, MIP, UR 4334, F-44000 Nantes, France

**Keywords:** Gait analysis, conventional gait model, segmental pose, marker set, inverse kinematics

## Abstract

**Background:** Three-dimensional gait analysis (3DGA) is widely used to support clinical decision-making in individuals with motor impairments. However, kinematic outputs depend strongly on the underlying biomechanical model. The open-source Conventional Gait Model II (CGM2) integrates updates to joint centre estimation (CGM2.1), inverse kinematics (CGM2.2), and cluster-based segment tracking (CGM2.3). While previous work demonstrated consistency among CGM2 variants in typically developing children, their effect in clinical populations remains unknown. This study quantified how CGM2 variants influence gait kinematics in individuals with cerebral palsy (CP).

**Methods:** Twenty-one individuals with CP (GMFCS I–II) underwent 3DGA using a 12-camera motion capture system and a CGM2.3 marker set. Hip, knee, and ankle kinematics from 487 gait cycles were computed using pyCGM2. Differences between CGM2.1, CGM2.2, and CGM2.3 were evaluated using Mean Absolute Deviation (MAD) and the adjusted coefficient of determination (R^2^).

**Results:** Overall, small differences were observed between model variants. MAD values were typically below 5° for most joints and planes, with high correlation between curves (R^2^>0.7). Hip rotation showed the largest discrepancies, with maximum MAD up to 7.7° when comparing CGM2.2 and CGM2.3. Differences between CGM2.1 and CGM2.3 were greater in the transverse and frontal planes but remained within acceptable limits (<5°), except for hip rotation.

**Conclusion:** The CGM2 variant selection has limited impact on gait kinematics in individuals with CP, and most differences fall within known repeatability error. However, transverse-plane kinematics, particularly hip rotation, should be interpreted with caution when comparing data across CGM2 variants.

## 1 Introduction

Three-dimensional gait analysis (3DGA) is a fundamental exam in the evaluation of human movement and supports clinical decision-making in individuals with motor impairments [1]. Its primary output, joint kinematics, is strongly dependent on the underlying biomechanical model and the marker-based processing pipeline [2]. The Conventional Gait Model (CGM) and its open-source successors, i.e. the Conventional Gait Model II (CGM2), are now widely used in 3DGA laboratories [3]. Successive CGM2 introduced key methodological refinements, including an updated hip joint centre estimation (CGM2.1) [4], kinematic-fitting based pose estimation (i.e., inverse kinematics) (CGM2.2) [5], and cluster-based segment tracking to compensate soft-tissue artefacts (CGM2.3) [6,7]. These developments address known limitations of the original CGM and aim to harmonize processing practices across laboratories [3].

In their recent work, Kerr et al.(2025) quantified the effects of CGM2 variants on gait kinematics and kinetics in typically developing (TD) children [8]. They reported that variants influence the transverse-plane parameters primarily, while the Gait Profile Score (GPS) remain consistent [8]. While this provided first evidence supporting the consistency of CGM2 within normative paediatric datasets, findings cannot be extrapolated to clinical populations. Individuals with cerebral palsy (CP) may present altered joint alignment [9] and pathological movement patterns [1] that may interact with model assumptions in non-linear and potentially amplifying ways.

Therefore, this study aims to quantify the impact of each CGM2 variant on kinematics in individuals with CP. This knowledge is essential both for research comparability across centres and for ensuring clinical decision-making is not confounded by model-dependent artefacts.

## 2 Materials and Methods

### 2.1.1 Participants

Twenty-one subjects with cerebral palsy (CP) (GMFCS I: n=17, GMFCS II: n=3, GMFCS III: n=1, age: 18.2(9.1) years old, height: 1.57(0.16) m, weight: 50.1(18.2) kg, and body mass index: 20.1(4.3) kg.m^-2^) were recruited. Informed consent was obtained, and the protocol conformed to the Declaration of Helsinki and was approved by the Institutional Review Board (“Commission Cantonale d’Éthique de la Recherche de Genève” (CCER-2020-00358)).

### 2.1.2 Data collection

The 3D trajectories of 23 skin-mounted and 4 wand-mounted light-reflective markers were recorded using a 12-camera optoelectronic system (Oqus7+, Qualisys, Sweden) sampled at 100 Hz. F**igure 1** details the location of the markers. Markers were placed according to the CGM markerset CGM2.3 [3,10] and their trajectories were filtered with a Butterworth fourth order with a cutting frequency of 6 Hz. All measurements were undertaken by the same assessor, with several years of experience in conducting 3DGA.

**Figure 1.**
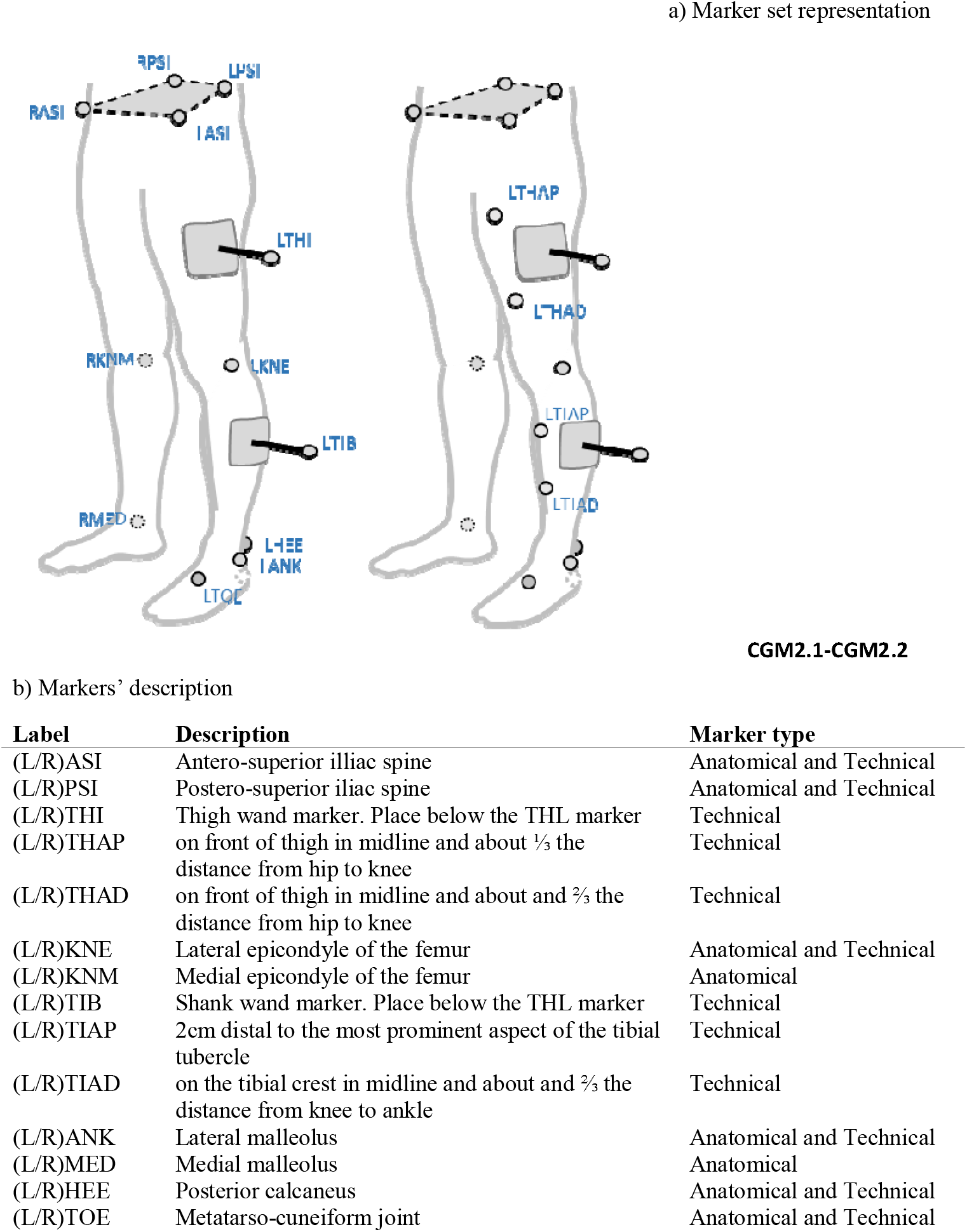
Marker set representation (a) and description (b).

### 2.1.3 Model processing

Knee and ankle joint centres were calibrated as the mid-point between the lateral and medial epicondyles or malleoli markers, respectively. The hip joint centres were calculated according to the predictive equations of Hara [11]. No correction of the knee varus-valgus was applied. These anatomical definitions were the same for all model variants (CGM2.1, CGM2.2, CGM2.3). For both CGM2.2 and CGM2.3, pose estimation was performed using kinematics fitting, with each joint modelled with 3 degrees of freedom (3dof).

### 2.1.4 Data processing

All computations were performed with the open-source package pyCGM2 [3]. Gait events were detected using force plate foot contacts and visually checked. A total of 487 cycles of the most impaired limb were analysed.

Overall comparisons of angles from each model were evaluated through the Mean Absolute Deviation (MAD) calculated as follows from a gait cycle:

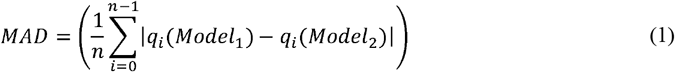

where *i* is the gait cycle timing, ranging from 1 to *n, q* stands for angles and models 1 and 2 represent two distinct model variants (CGM2.1 or CGM2.2 or CGM2.3).

Curve similarity was quantified with the adjusted R^2^ [12]. Interpretation of the coefficient followed rules [26]: Strong correlation (R^2^>0.9); high (0.7<R^2^≤0.9); moderate (0.5≤R^2^<0.7), low (0.3≤R^2^<0.5), negligible (R^2^<0.3)

## 3 Results

Gait kinematics from the different model variants is presented in **Figure 2**. The hip transverse rotation presented the largest MAD (mean MAD (standard deviation)–maximum = CGM2.1 vs CGM2.2: 2.6(0.8)–4.2; CGM2.1 vs CGM2.3: 4.1(1.0)–5.9; CGM2.2 vs CGM2.3: 3.1(1.7)–7.7 (**Table 2**).

**Figure 2.**
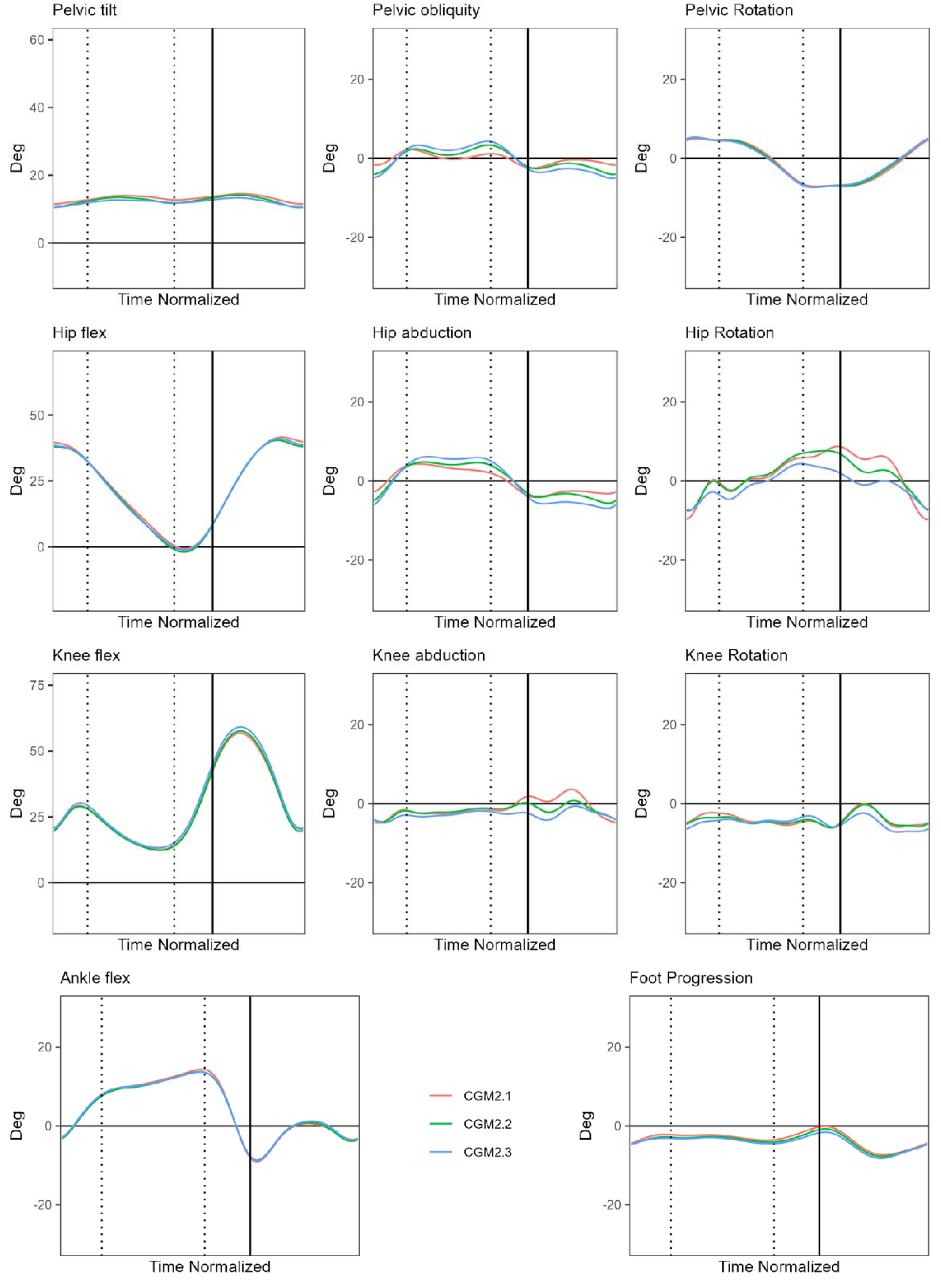
Gait kinematics from the different model variants. The mean curve for the entire CP cohort is presented.

The mean correlation between each model variant was high for all kinematics parameters (R^2^>0.7), except for the pelvis obliquity between CGM2.1 and CGM2.3: mean (standard deviation): 0.6(0.3) (**Table 2**).

## 4 Discussion

Overall, the effect of CGM2 variants on gait kinematics of individuals with CP was small. For most joints and planes, the MAD remained below 5°, with good waveform agreement (R^2^>0.7).

Differences between CGM2.1 and CGM2.2 variants arise from inverse kinematics optimally fitting the model to experimental marker data under joint-level constraints, potentially reducing sensitivity to soft-tissue artefacts [13]. Results show small MAD (maximum MAD<5°) and good correlation (R^2^>0.7) for all joint kinematics, consistent with Kainz et al. (2016), who reported errors less than 5° for all joint angles between inverse and direct kinematics models when the models shared the same definition of segmental references and marker count [2]. Similar differences were reported by Kerr et al. (2025), comparing CGM2.1 and CGM2.2 resulting kinematics in TD individuals, with no significant difference in the GPS [8]. Conversely, Hayford et al. (2021) observed GPS changes with kinematic fitting, although clinical interpretation remained unaffected [14]; however, they did not model all joints with 3dof [14], unlike the CGM2.2.

Differences between CGM2.2 and CGM2.3 are attributable to marker set changes, with thigh and shank segments tracked using three markers (CGM2.3) instead of a single lateral marker (CGM2.2). Hip rotation showed a larger difference (mean MAD=3.1° – max 7.7°), although mean correlation remained high (R^2^=0.8). These results are consistent with previous studies that reported that marker-set changes lead to observable discrepancies in kinematics, especially for transverse rotations [15,16].

The largest differences were observed between the CGM2.1 and CGM2.3, particularly for the transverse and frontal planes. Except for hip rotation, differences remained below the acceptable 5° threshold [17], and typically repeatability error, unlikely to alter clinical interpretation. However, hip rotation results consistently exceeded the 5° threshold and should therefore be interpreted with caution [17]. It is well established in the gait analysis literature that kinematic measurements in the transverse and frontal planes are more susceptible to measurement error [17], compared to the sagittal plane [15,16,18–20].

All analyses used the open-source library pyCGM2 [3], embedding the OpenSim API [12] for kinematic fitting. All CGM2 variants shared the same anatomical definitions, ensuring consistent joint and segment representation. Without such consistency, differences could be substantially larger, as illustrated by Ziziene et al. (2022), who reported a mean difference across all lower limb joints of 13°[21].

Several limitations should be acknowledged. First, no gold standard for the true bone orientation was used. Results reflect between-model differences rather than trueness. Second, analyses were restricted to individuals with GMFCS level I predominantly, who present with mild impairments. Further investigations may be undertaken in individuals with more severe or other musculoskeletal disorders. Third, kinetic results were not reported; given small kinematic differences and a common anthropometric model for inverse dynamics within pyCGM2, joint moments were not expected to be substantially affected. Our study focused on kinematics, the most widely accessible and interpretable outputs in clinical gait analysis.

In conclusion, CGM2 variant selection has limited impact on gait kinematics in individuals with CP. However, clinicians and researchers should interpret transverse-plane outputs with caution when comparing data from different CGM2 variants. Using the CGM 2.3 marker set as the preferred systematic standard provides the flexibility to leverage earlier model variants (CGM 2.1 and CGM 2.2) when needed, while simultaneously fostering the adoption of musculoskeletal modelling through a consistent, robust, and forward-compatible biomechanical framework.

## Data Availability

All data produced in the present study are available upon reasonable request to the authors

---

**Table 1.** Comparisons between each model variant. For each comparison, the mean absolute deviation (MAD), standard deviation (sd), and maximum are presented, as well as the mean R^2^ and sd. MAD over the acceptable 5° threshold (maximum) are bold.

|  |  |  | CGM2.2 |  | CGM2.3 |  |
| --- | --- | --- | --- | --- | --- | --- |
| Parameters |  |  | MAD<br>Mean (sd) -<br>maximum | R<sup>2</sup><br>Mean (sd) | MAD<br>Mean (sd) -<br>maximum | R<sup>2</sup><br>Mean (sd) |
| CGM 2.1 | Pelvis | Tilt | 0.7 (0.6) - 2.2 | 0.9 (0.1) | 1.1 (0.7) - 2.6 | 0.9 (0.1) |
|  |  | Obliquity | 1.5 (0.5) - 2.3 | 0.7 (0.2) | 2.4 (0.8) - 3.6 | 0.6 (0.3) |
|  |  | Rotation | 0.5 (0.2) - 0.8 | 1.0 (0.0) | 0.6 (0.2) - 1.0 | 1.0 (0.0) |
|  | Hip | Flexion | 1.2 (0.6) - 2.7 | 1.0 (0.0) | 1.3 (0.4) - 2.4 | 1.0 (0.2) |
|  |  | Abduction | 1.5 (0.5) - 2.4 | 0.9 (0.1) | 2.8 (0.9) - 4.1 | 0.8 (0.1) |
|  |  | Rotation | 2.6 (0.8) - 4.2 | 0.9 (0.1) | **4.1 (1.0) - 5.9** | 0.7 (0.1) |
|  | Knee | Flexion | 1.1 (0.5) - 2.1 | 1.0 (0.0) | 1.7 (0.6) - 2.8 | 1.0 (0.0) |
|  |  | Abduction | 1.7 (0.8) - 3.2 | 0.9 (0.1) | 2.1 (0.9) - 4.4 | 0.7 (0.2) |
|  |  | Rotation | 1.0 (0.5) - 2.5 | 1.0 (0.0) | 2.0 (0.5) - 2.8 | 0.8 (0.1) |
|  | Ankle | Flexion | 0.9 (0.4) - 2.0 | 1.0 (0.0) | 1.0 (0.6) - 2.8 | 1.0 (0.0) |
|  | Foot | Progression | 0.6 (0.2) - 1.0 | 1.0 (0.0) | 0.9 (0.5) - 1.8 | 1.0 (0.0) |
| CGM 2.2 | Pelvis | Tilt |  |  | 0.5 (0.2) - 0.9 | 0.9 (0.1) |
|  |  | Obliquity |  |  | 1.1 (0.3) - 1.4 | 0.9 (0.1) |
|  |  | Rotation |  |  | 0.3 (0.1) - 0.5 | 1.0 (0.0) |
|  | Hip | Flexion |  |  | 0.8 (0.2) - 1.3 | 1.0 (0.0) |
|  |  | Abduction |  |  | 1.5 (0.5) - 2.4 | 1.0 (0.0) |
|  |  | Rotation |  |  | **3.1 (1.7) - 7.7** | 0.8 (0.2) |
|  | Knee | Flexion |  |  | 1.4 (0.5) - 2.4 | 1.0 (0.0) |
|  |  | Abduction |  |  | 1.3 (1.1) - 4.3 | 0.9 (0.1) |
|  |  | Rotation |  |  | 1.6 (0.3) - 1.8 | 0.9 (0.1) |
|  | Ankle | Flexion |  |  | 0.4 (0.2) - 0.6 | 1.0 (0.0) |
|  | Foot | Progression |  |  | 0.4 (0.1) - 0.9 | 1.0 (0.0) |

